# RapiD_AI: A framework for Rapidly Deployable AI for novel disease & pandemic preparedness

**DOI:** 10.1101/2022.08.09.22278600

**Authors:** Alexey Youssef, Tingting Zhu, Anshul Thakur, Peter Watkinson, Peter Horby, David W Eyre, David A Clifton

## Abstract

COVID-19 is unlikely to be the last pandemic that we face. According to an analysis of a global dataset of historical pandemics from 1600 to the present, the risk of a COVID-like pandemic has been estimated as 2.63% annually or a 38% lifetime probability. This rate may double over the coming decades. While we may be unable to prevent future pandemics, we can reduce their impact by investing in preparedness. In this study, we propose *RapiD_AI* : a framework to guide the use of pretrained neural network models as a pandemic preparedness tool to enable healthcare system resilience and effective use of ML during future pandemics. The RapiD_AI framework allows us to build high-performing ML models using data collected in the first weeks of the pandemic and provides an approach to adapt the models to the local populations and healthcare needs. The motivation is to enable healthcare systems to overcome data limitations that prevent the development of effective ML in the context of novel diseases. We digitally recreated the first 20 weeks of the COVID-19 pandemic and experimentally demonstrated the RapiD_AI framework using domain adaptation and inductive transfer. We (i) pretrain two neural network models (Deep Neural Network and TabNet) on a large Electronic Health Records dataset representative of a general in-patient population in Oxford, UK, (ii) fine-tune using data from the first weeks of the pandemic, and (iii) simulate local deployment by testing the performance of the models on a held-out test dataset of COVID-19 patients. Our approach has demonstrated an average relative/absolute gain of 4.92/4.21% AUC compared to an XGBoost benchmark model trained on COVID-19 data only. Moreover, we show our ability to identify the most useful historical pretraining samples through clustering and to expand the task of deployed models through inductive transfer to meet the emerging needs of a healthcare system without access to large historical pretraining datasets.

## 1 Introduction

COVID-19 has introduced a shock that has substantially disrupted often unprepared healthcare systems globally and exposed an array of gaps and vulnerabilities that contributed to the sub-optimal outcomes during the pandemic [1, 2]. Haldane and colleagues [1] developed a framework of healthcare system resilience based on the WHO building blocks of healthcare systems: (1) Governance, financing, and collaborations across sectors, (2) Community engagement, (3) Health Service Delivery, (4) Health workforce, (5) Medical Products and technologies, and (6) Public health functions. Resilient healthcare systems are identified by their ability to (A) Activate comprehensive responses, (B) Adapt health system capacity, (C) Preserve health system functions and resources, and (D) Reduce vulnerability. Such systems are robust to external shocks and are proactive rather than reactive in managing and mitigating risk with heavy investment in preparedness and the ability to scale response to threats effectively when challenged. Capacity and resilience during the COVID-19 pandemic were frequently inadequate. The pandemic illustrated that the current metrics used to measure the preparedness of healthcare systems should change as they did not reflect performance during the pandemic. For example, the US was ranked atop the Global Health Security Index, yet it was one of the hardest-hit countries [3]. There is expert consensus that we will have an increasing number of pandemics in the future as our world becomes more connected. The latest estimates illustrate that the rate of extreme pandemics is higher than previously believed and that it is accelerating: 2.63% annually or a 38% lifetime probability, with the possibility to double in the coming decades [4]. In the context of pandemic preparedness, relatively little attention is channelled towards data science and AI. Healthcare systems face tremendous pressures during a novel disease pandemic. AI has the potential to optimise and assist the performance of healthcare systems in many areas (pandemic spread prediction, infectious agent mutation prediction, patient triage, patient management, and diagnosis). The COVID-19 pandemic is the first pandemic that witnessed a proliferation of machine learning solutions and offers us an opportunity to learn about the successes and challenges of machine learning implementation [5–10]. Some of the challenges are below

1. Machine learning applications during the pandemic were reactive due to the lack of investments in machine learning preparedness.
2. The lack of access to data was a major barrier to building machine learning algorithms. Two factors contribute to the lack of data (i) COVID-19 is caused by a new infectious agent that healthcare systems have not seen before. This means that historical disease data is unavailable, and (ii) the lack of national and regional frameworks that allow effective data pooling and aggregation across healthcare facilities means that data ends up in small silos too small to enable the train of effective AI algorithms.
3. Distributional shifts (e.g., incidence changes, outcome prevalence, population characteristics) occur during the different phases of the pandemic. These may be driven by population-level control measures, behaviour and healthcare use changes, and the impact of novel pathogen variants, vaccinations, and treatments. Distribution shifts substantially impact machine learning performance, especially when training data is limited. This creates variability in model performance, lack of robustness, bias, and many other challenges. Singh and colleagues [11] demonstrate the impact of distribution shifts across 179 US facilities on a sepsis prediction model. Duckworth and colleagues [12] show distribution shifts during the COVID-19 pandemic and evaluate their impact on a hospital admission prediction model.

Taking the above challenges into account, the effective utilisation of AI in the context of a novel disease pandemic will require a shift towards a proactive development and implementation framework. The standard ML development framework includes a phase of data collection or data acquisition. This is especially important in problems on which we do not have enough historical data, such as a novel disease. In the context of a pandemic, human life is too precious to wait to collect enough data to train and deploy AI. Therefore, we need better development approaches that overcome the limited availability of data during pandemics. We present the concept of Rapidly Deployable AI models (RapiD_AI) as a novel disease and pandemic preparedness tool. The RapiD_AI framework enables healthcare systems to leverage the availability of historical non-pandemic data to build models that can be rapidly deployed in the case of a novel disease using small amounts of data collected during the early encounters of the healthcare system with the novel pathogen [figure 1]. We provide an experimental demonstration of the framework by comparing two sets of models. The first is ***pretrained*** : models trained historical patient data and fine-tuned using COVID-19 data collected during the first weeks of the pandemic. The second is ***trained from scratch*** : models trained on the COVID-19 data collected during the first weeks of the pandemic. We show that we can (i) use pretraining to gain a performance advantage over training from scratch [Scenario A], (ii) use a clustering approach to select the most useful pretraining samples to produce non-inferior performance [Scenario B], and (iii) expand the task of a deployed pretrained model to a pandemic-related task using inductive transfer [Scenario C] [figure 2].

**Fig. 1.**
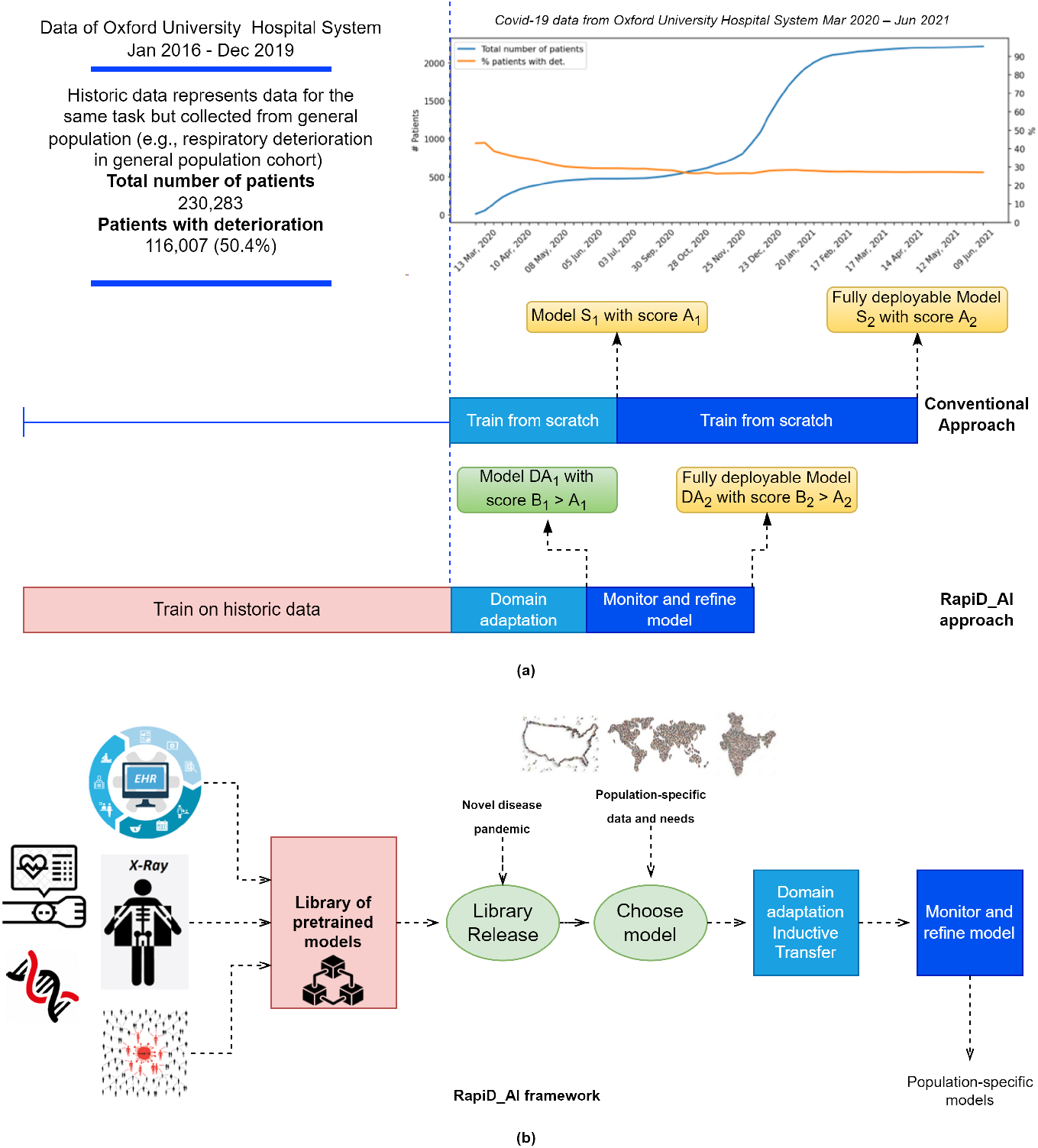
Section (a) provides an overview of the COVID-19 and historical training data used for our experimental workflow, along with a timeline to demonstrate the training and deployment process. Section (b) shows the work schema RapiD_AI framework for the development of rapidly deployable AI models as a pandemic preparedness tool: (1) Different sources of data can be used to pretrain many models (2) During new pandemics, these models can be released (3) These models can be fine-tuned using limited population-specific local data gathered in the first weeks of the pandemic to generate population-specific models across a variety of tasks and data categories (e.g., EHR tabular, imaging, genomics data, etc.). Such an approach addresses the issue of limited data available in the context of novel disease pandemics

**Fig. 2.**
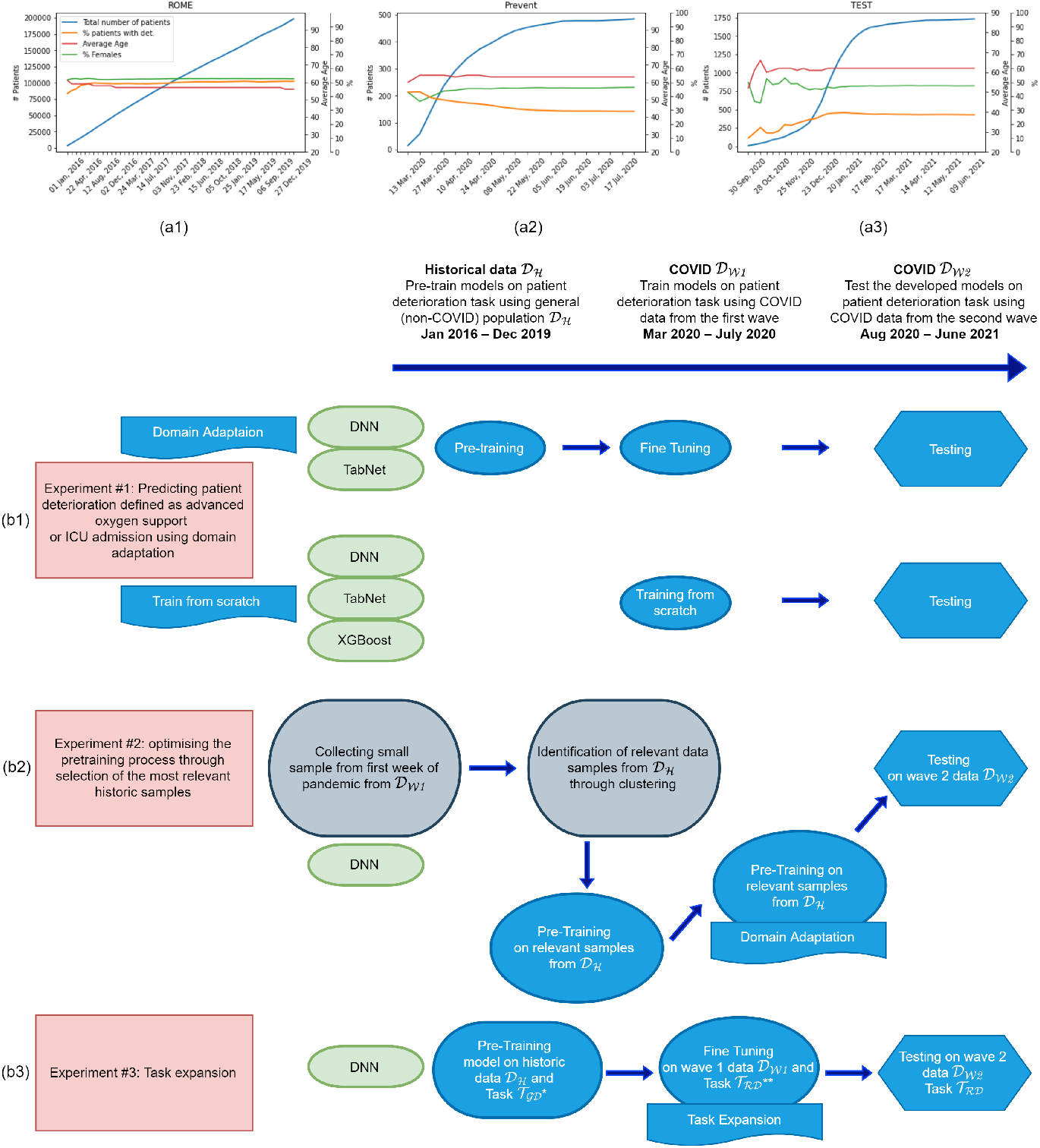
Sections (a1-a3) shows basic statistics (Total number of patients, percentage of patients with deterioration, average age, and percentage of females) of our datasets 𝒟_*H*_, 𝒟_*W1*_, and 𝒟_*W2*_ respectively. Sections (b1-b3) show each dataset’s use in our 3 experimental workflows. We used three models in our experiments: Deep Neural Network (DNN), TabNet, and XGBoost. Our neural network models (DNN & TabNet) are pretrained on 𝒟_*H*_ and fine-tuned on weekly cumulative data from 𝒟_*W1*_, while XGBoost, DNN (from scratch) and TabNet (from scratch) models are trained with weekly cumulative data from 𝒟_*W1*_. All models are tested on 𝒟_*W2*_ except for DNN and XGBoost from figure 4 (c) where test data is a smaller segment of 𝒟_*W2*_ given we extended 𝒟_*W1*_ and reduced the size of 𝒟_*W2*_. *𝒯_ℛ𝒟_ is the respiratory deterioration task, **𝒯_𝒢𝒟_ is the general deterioration task. (A detailed description is available in the results & methods sections)

## 2 Results

### 2.1 Dataset

In our work, we used three datasets extracted from the same population:

1. 𝒟_*H*_: Pre-pandemic dataset from Jan 2016 to Dec 2019. General in-patient cohort.
2. 𝒟_*W1*_: March 2020 to July 2020. COVID-19 patients only. This represents the first wave of the pandemic.
3. 𝒟_*W2*_: August 2020 to June 2021. COVID-19 patients only. This represents the second wave of the pandemic.

Each patient in the dataset has multiple observations, each of them characterised by a 77-dimensional feature vector and a label exhibiting the occurrence of a respiratory deterioration event within the next 24 hours. The features include commonly measured vital signs and laboratory parameters as well as the variability of those features over time D4. Figure 2 (a1 & a2 & a3) illustrates the cumulative number of patients as well as the demographic statistics of patients and the rate of deterioration for the three datasets: 𝒟_*H*_, 𝒟_*W1*_, 𝒟_*W2*_.

### 2.2 Task Definition

In our experimental process, we use two distinct task definitions. Both are composite *patient deterioration prediction* tasks. The first is a ***respiratory deterioration prediction task* 𝒯**_***RD***_, a composite task based on the escalation in the level of oxygen support requirement in a forward-looking window of 24 hours from level 0/1 to level 2/3 or unplanned ICU admission (as a proxy for mechanical ventilation in COVID-19 patients). We defined the level of support based on the oxygen delivery devices used. A list of the devices and the corresponding level of support is documented in A1. The second is a ***general deterioration prediction task* 𝒯**_***GD***_, defined as the composite outcome of mortality or ICU admission in a forward-looking window of 24 hours. In both tasks, outcomes happening less than 1 hour after admission were excluded. In the case of multiple occurrences of adverse events for one patient, we removed observations recorded after the first event.

### 2.3 Experimental setup

Our experimental setup is based on three scenarios (A, B, C). An XGBoost model trained from scratch on weekly COVID-19 data was used as the benchmark performance measure in the three scenarios. We chose XGBoost because it is one of the standard go-to models for tabular data problems [13]. Moreover, XGBoost has demonstrated strong performance on tasks similar to those used in our experimental workflow. The PreVent study compared various ML models to predict respiratory deterioration in COVID-19 patients and demonstrated that XGBoost achieved the best performance [14].

#### 2.3.1 Scenario A: Domain adaptation

In this experimental section, we sought to illustrate the value of using historical data in pretraining RapiD_AI models in the context of a pandemic of a novel disease [figure 2 (c)]. We have used 𝒟_*H*_ to pretrain our deep learning models. We used 𝒟_*W1*_ to either (i) train our benchmark XGBoost and neural network models (DNN & TabNet) from scratch or (ii) fine-tune the pretrained networks. We used 𝒟_*W2*_ as a held-out test dataset to evaluate the performance of the models.

We chose Deep Neural Network and TabNet models to test the RapiD_AI framework. TabNet is a deep learning sequential attention-based model released by Google Brain. Its authors argue that the model has the potential to dominate tabular data problems [15]. Our benchmark model is an XGBoost model trained on cumulative weekly data from 𝒟_*W1*_.

##### Model performance

Our results show that pretraining deep neural models boosted their performance over the first 20 weeks of the pandemic. Pretraining improved DNN performance by relative/absolute 110.87/41.71% AUC in the first week of the pandemic and then by a stable absolute average of 3.86% AUC over the next 19 weeks [figure 4 (b1)]. Pretraining improved TabNet performance by an average relative/absolute gain of 16.24/9.16% AUC over the first 20 weeks of the pandemic [figure 4 (b2)].

**Fig. 3.**
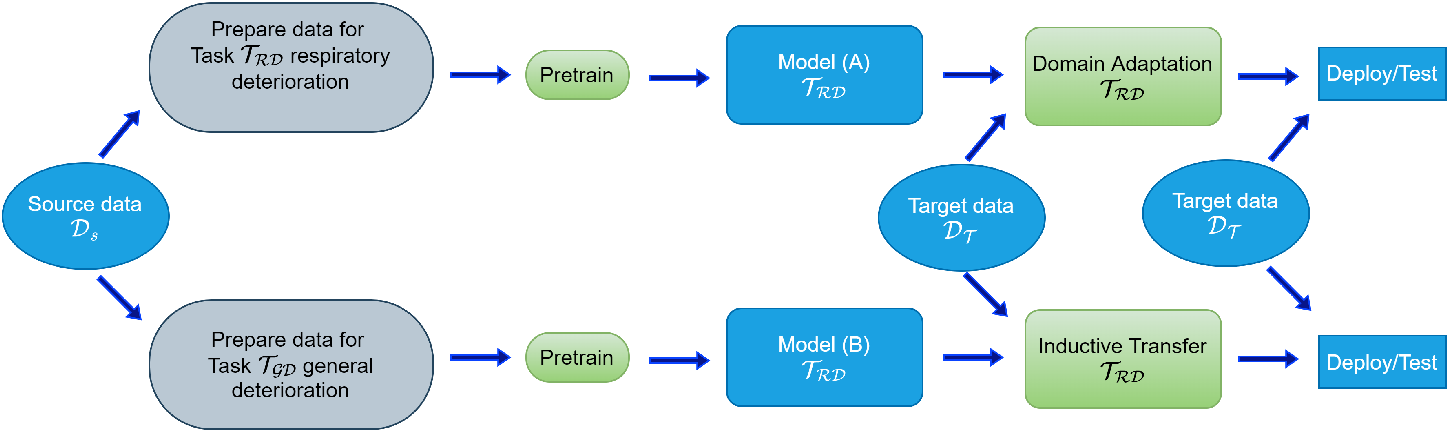
The difference between two transfer learning approaches used in our work (domain adaptation and inductive transfer). In domain adaptation, we have the same task but different data distribution in source and target domains. In inductive transfer, the source and target domain tasks differ, while the data distribution could be the same or different. 𝒟_𝒮_ is the source data from which we extracted pretraining datasets, 𝒟_𝒯_ is the target domain data. 𝒯_ℛ𝒟_ is the Respiratory deterioration task. 𝒯_𝒢𝒟_ is the general deterioration task. (A detailed description is available in the results & methods sections)

**Fig. 4.**
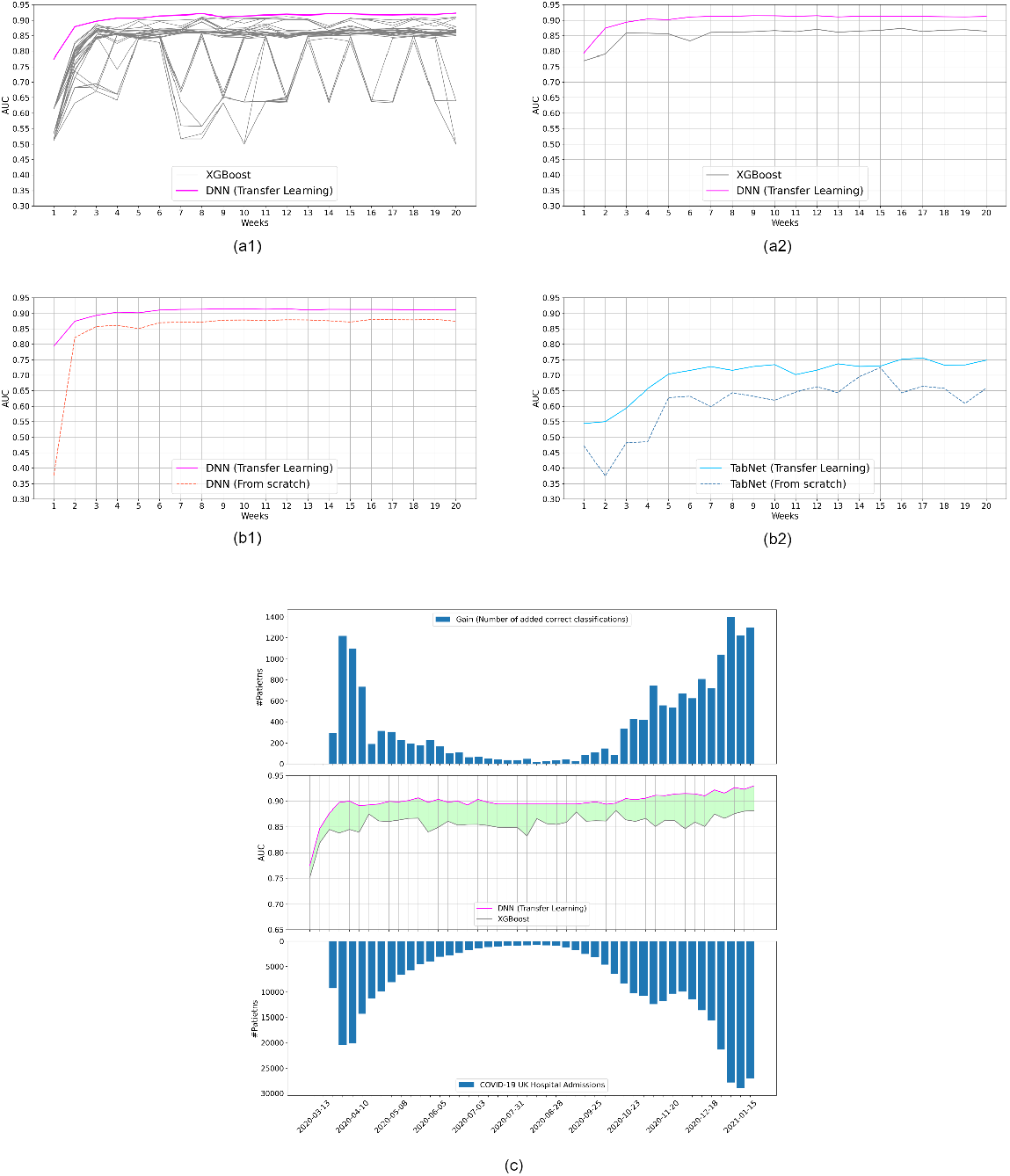
Section (a1) compares the average performance of DNN (transfer learning) with many configurations of XGBoost based on different sets of hyperparameters. Using the best configuration of the first three weeks, we plot in (a2) the average performance and confidence intervals of XGBoost and DNN (transfer learning) models. Section (b1) shows the difference in DNN performance with and without pretraining. Section (b2) shows the same as (b1) but for the TabNet. The lower part of (c) shows the weekly number of COVID-19 hospital admissions in the UK between March 2020 and Jan 2021; the middle part shows the difference in AUC between XGBoost and pretrained DNN and the upper part reflects an attempt at measuring the potential clinical performance gain. Gain is measured by scaling the algorithmic AUC performance gain (as a %) by a factor of COVID-19 weekly hospital admissions in the UK. The algorithmic gain is measured as the AUC performance differential between the pretrained DNN and the benchmark XGBoost. This is based on the assumption that an algorithm is triggered on average once per admission. The number illustrates the improved performance delivered by a pretrained DNN compared to an XGBoost trained on COVID-19 data only.

We also extended the evaluation window to include the first ten months of the pandemic (Mar ‘20 - Jan 21). We tested our RapiD_AI DNN model against baseline XGBoost trained on the cumulative weekly data [figure 4 (c)]. We show that our RapiD_AI universally outperforms the baseline XGBoost model by relative/absolute 4.37/3.58% AUC in the first four weeks of the pandemic and relative/absolute 4.92/4.21% AUC on average in the whole period. In the case of a novel global disease pandemic, these small gains in performance can potentially translate into significant clinical and operational benefits. In the first 23 months of the pandemic (until January 2022) the UK experienced 678,091 COVID-19 hospital admissions [16]. The lower section of figure 4 (c) shows the number of weekly COVID-19 cases in the UK, and the upper part reflects the performance gain we can achieve. The clinical impact is measured by scaling the performance gain (as a %) by a factor of COVID-19 weekly hospital admissions in the UK. The performance gain is measured as the AUC performance differential between the pretrained DNN and the benchmark XGBoost. The 4.21% average gain in algorithm AUC translates into a gain of up to 1,399 added correct classifications weekly if rolled across the UK National Health Service. This number could translate into better targeted medical interventions for thousands of patients.

##### Model robustness

Our results demonstrate that RapiD_AI models are more robust than baseline XGBoost and non-pretrained models. Figure 5 (a1) illustrates the variability in the performance of the XGBoost model with different hyperparameter (HP) sets. With certain sets of hyperparameters, the model performs well in some weeks and poorly in others. For example, using the first three months of our training data, we still get fluctuating AUC (63%-91%) depending on our chosen hyperparameters. Generally, there is no universally best-performing HP set, and achieving the optimal performance of the model would require frequent HP fine-tuning if implemented in practice. Moreover, figure 4 (a2) illustrated that our RapiD_AI DNN has consistently narrower Confidence Interval CI compared to a baseline XGBoost over the first 20 weeks of the pandemic. Figures 5 (b1 & b2) illustrate that pretrained DNN and TabNet models have narrower CI compared to the same models trained from scratch on weekly cumulative data. This consistency would translate into more stable performance after deployment. Figure 5 (c) demonstrates the RapiD_AI DNN stability and superiority over time compared to XGBoost.

**Fig. 5.**
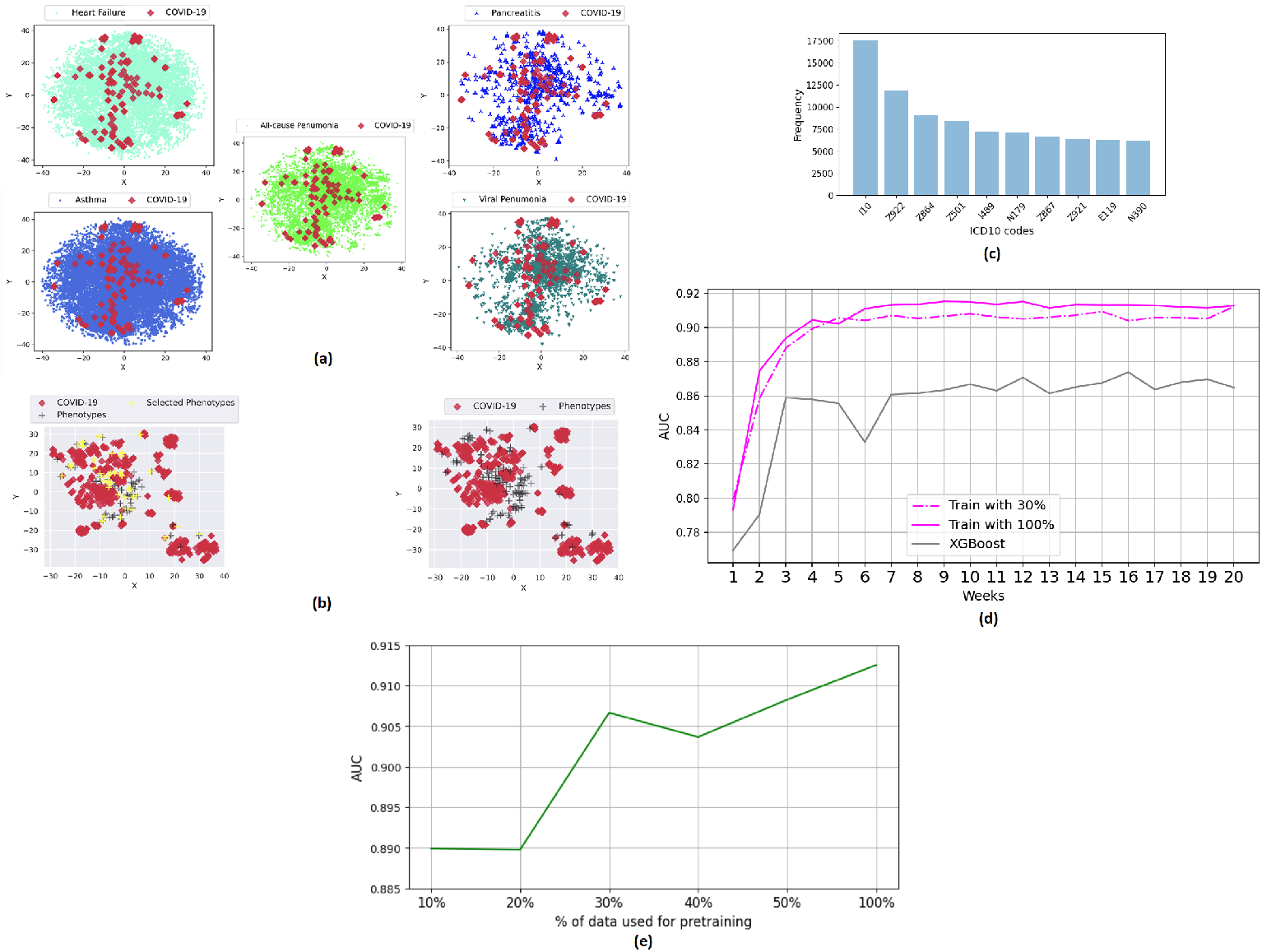
The figure outlines the results of the clustering experiments to identify the most relevant historical pertaining examples. Section (a) shows the similarity between COVID-19 samples and five selected disease populations based on ICD-10 codes. Of the five groups, COVID-19 patients are most similar to Acute Pancreatitis and Viral Pneumonia patients. Section (b) plots COVID-19 samples against cluster centres of the historical pretraining data. Based on this plot, we select the most similar 10, 20, 30, 40, and 50% of historical data clusters and use the selected data for pretraining. Section (c) shows the most frequent ICD-10 codes in the top 10% of the selected clusters. Section (d) Shows that a pretrained DNN model with only 30% of data can result in non-inferior performance compared to a model pretrained on 100% of historical data. Section (e) shows the performance of our DNN model as a function of the proportion of data used for pretraining. AUC was averaged over the last five weeks of 𝒟_*W1*_ dataset.

#### 2.3.2 Scenario B: Pretraining optimisation

This experiment aims to replicate the scenario of a healthcare system facing a novel disease pandemic while having access to sufficient historical data. The source domain (pretraining historical data) is large (more than nine million samples) and diverse. It represents all possible clinical conditions that require an in-patient admission within the source population. In this scenario, we hypothesized that by selecting the most relevant pretraining examples, we might achieve superior performance compared to pretraining on all historical data while reducing the computational demand of the pretraining process. We considered the historical examples with maximum similarity to the COVID-19 data to be the most relevant. The selection of pretraining samples was attempted in two ways:

- Using human experience: We asked a physician with COVID-19 care experience to identify five disease clusters with various degrees of similarity to the observed clinical pattern of COVID-19 in the first week of the pandemic. We identified five clusters with various degrees of similarity: viral pneumonia (similar), all-cause pneumonia (similar), acute pancreatitis (dissimilar), Asthma (dissimilar), and heart failure (dissimilar). The ICD10 codes used to identify each condition are listed in B2. After that, we plotted the similarity of COVID-19 patients to these five clusters using a tSNE clustering approach [figure 5 (a)]. The clusters most similar are viral pneumonia (expected) and pancreatitis (unexpected). This indicates that, in our case, the training examples with the most similar distribution to COVID-19 (i.e., most useful training examples) can not be identified using clinical intuition alone.
- Using computational approach: We used a computational approach to identify the most useful training examples. Using tSNE, we plotted clusters of all the historical data along with the COVID-19 samples from the first three weeks [figure 5 (b)]. Using clustering, we identified historical clusters similar to COVID-19 and pretrained on these clusters. We did pretraining experiments with 10%, 20%, 30%, 40%, and 50% of data and found that using 30% & 50% of pretraining clusters resulted in the best balance. Figure 5 (e) shows the average performance over the last five weeks when using different data partitions. Figure 5 (d) shows a detailed performance comparison between 3 models: XGBoost, DNN model pretrained on 30% of data and DNN model pretrained on all available data. A DNN pretrained on 30% of the clusters demonstrated non-inferior performance compared to a network pretrained on all data.

We identified the most frequent ICD10 codes in the 10% most similar clusters [figure 5 (c)]. The top 10 codes were: I10, Z922, Z864, Z501, I489, N179, Z867, Z921, E119, N390. The full list of ICD-10 [appendix table B3]. We observe that the most frequent ICD10 codes were not clinically intuitive. However, this is compounded given (i) the frequency of the codes in the general population may influence the composition of the most frequent ICD10 codes in the selected training clusters, and (ii) multiple ICD10 codes per patient make it difficult to discern the code of primary diagnosis.

#### 2.3.3 Scenario C: Task expansion & inductive transfer

This scenario aims to replicate the scenario of a healthcare system facing a novel disease pandemic while having access to deployed machine learning models but not historical data. Using the approach of inductive transfer, we repurposed a pretrained deployed model to build an algorithm that would address a different clinical problem in the context of a novel pandemic. A pretrained DNN on a general patient deterioration task 𝒯_*GD*_ was fine-tuned using weekly COVID-19 data to perform a respiratory deterioration task 𝒯_*RD*_ in COVID-19 patients. The algorithm demonstrated a performance relative/absolute gain of 11.93/9.32% AUC over the first two weeks of the pandemic compared to an XGBoost trained only on weekly data. The performance gain was sustained over the first 20 weeks of the pandemic with an average AUC relative/absolute increase of 7.57/6.42%. Inductive transfer showed a dominant performance over domain adaptation (scenario A) and clustering (scenario B) [figure 6 (b)]. The dominant performance of the inductive transfer approach over domain adaptation can be attributed to the fact that the knowledge gained from training on two different tasks boosted the model capacity. This can be seen similar to the benefits multi-task learning introduces to the generalisation of models [17]. Besides the dominant performance, this model can be deployed faster than others as the pretrained part is assumed to be ready and borrowed from a previously trained task.

**Fig. 6.**
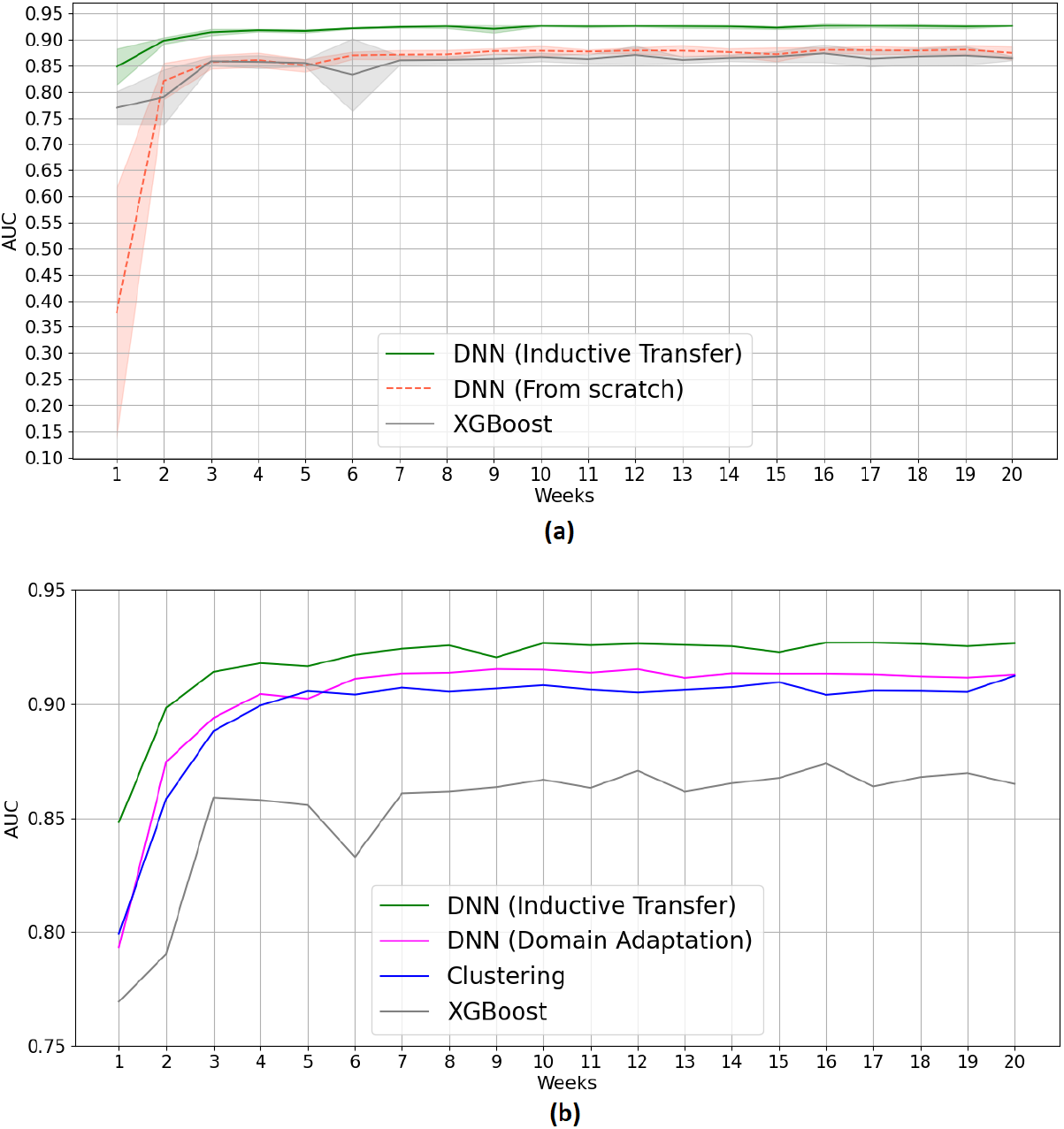
The figure summarizes the experimental results for scenario C. Section (a) shows the superior performance of inductive transfer over models trained from scratch. Section (b) compares the performance of models trained using inductive transfer and domain adaptation and the performance of models pretrained on 30% of data identified using clustering from scenario B. All the provided models showed superiority over the XGBoost model.

## 3 Conclusions

We show that pretrained DNN models can deliver superior performance during a novel disease pandemic compared to DNN and XGBoost models trained from scratch on cumulative pandemic data alone. We show gains in AUC, consistency, and robustness (measured by Confidence Interval and performance variability). Moreover, we demonstrate that we can identify the most relevant historical training examples. We use 30% of the pretraining data to produce a model that approximates the performance of a model pretrained on the entire historical dataset. We also show that by using inductive transfer, we readjust a deployed model to our COVID-19 task. Beyond those results, our work highlights a few key points:

First, methodologies are needed to address the lack of training data during a pandemic. During healthcare emergencies caused by a new pathogen or disease, healthcare systems are exposed to an increased demand by clinical cases with unique pathophysiology not described before. In the early phases, the lack of data on the novel infectious agent limits the ability to develop machine learning and statistical tools that can assist healthcare systems in coping with emergencies.

The lack of data exposes models to a high risk of bias and limits generalisability. Models developed in the early phase of the pandemic have indeed demonstrated methodological limitations like bias, lack of validation, and calibration [18–22]. Both domain adaptation and inductive transfer methodologies can address the lack of data. Yet, despite the promise and positive results of transfer learning, its applications in healthcare are very limited [23–25]. This work is one of the few demonstrations of transfer learning on tabular EHR data.

Second, machine learning algorithms are susceptible to distribution shifts. Modern AI and deep learning show excellent performance as long as the training and test data are from a similar population/distribution [26]. Hence, a model trained on one population may not perform when deployed on a different population, even for the same task. In that sense, Deep Learning is local [27, 28]. The local nature of Deep Learning calls for model domain adaptation and fine-tuning when deploying to populations different from the training set, allowing models to capture the distribution shifts that occur across populations due to workflow, genetic, environmental, and other differences. In the context of a pandemic, algorithms are also susceptible to distribution shifts due to the changing dynamics of the disease/population over time (e.g., vaccines have reduced the severity of COVID-19 cases). To capture these shifts, we should incorporate online and incremental learning (frequent model retraining) into the local deployment pipeline. Recent regulatory updates such as the FDA algorithmic predetermined change control plan make the localisation and frequent fine-tuning practically, and regulatorily possible [29].

Third, we demonstrate that while the computational efficiency of the pretraining process can be optimised, selecting the most relevant pretraining samples does not lead to superior performance in our case. We also show that identifying the most useful training examples is not clinically intuitive. Hence an optimal pretraining optimisation approach is likely to be either purely computational or hybrid computational-clinical. This raises the question of whether machines identify clinical patterns that humans overlook. One potential reason is that humans and machines have different representations of diseases. Humans understand diseases based on the underlying pathophysiological processes. Machines see data only and understand diseases based on data. From a clinical standpoint, COVID-19 acute pneumonia and acute pancreatitis are very different diseases. From a machine’s perspective, these two disease can be similar in the acute deterioration and decompensation patterns.

In this paper, we showcase the RapiD_AI framework as a pandemic preparedness tool and a conceptual framework for the implementation of machine learning during a novel disease pandemic [figure 1 (b)]. To overcome the limitations brought forward by the lack of data and pathogen novelty, we suggested using historical data to pretrain rapidly deployable models. The models can be fine-tuned to reach optimal performance rapidly using small amounts of data collected during the early encounters with the novel pathogen at a local level.

RapiD_AI framework suggests building rapidly deployable neural network models using historical data to circumvent the need for large quantities of novel disease training data. The models can be pretrained on historical data from different domains such as biomedical signals (e.g., ECG), EHR, medical imaging, genomics, and epidemiology [figure 1 (b)]. This approach leads to a library of models specific to different data modalities: an ECG model, an EHR model, a CT model, etc. These models can be rapidly deployed when the need emerges to support the pandemic response. The models can address predictive tasks that vary according to the data modality (e.g., patient triage and deterioration prediction for EHR models or disease spread prediction for epidemiological models). The models can be built using global data, maintained by health authorities, and released in the event of a pandemic. Local healthcare stakeholders can (i) use the data they collect during the first weeks of the pandemic to fine-tune the pretrained models using domain adaptation or inductive transfer, and (ii) maintain these local models as the pandemic unfolds using online learning and fine-tuning. This approach uses generic pretrained deep learning models to develop localised versions of the model that address the local needs of healthcare facilities and populations either by allowing the adaptation of the model to the local patient data distribution or by adapting the predictive task according to the needs of the local facilities.

While it is true that historical data can be different than the disease at hand, pretraining is helpful because neural networks benefit from being exposed to a large quantity of data similar in structure to the data of the target task. A large pretraining dataset helps the neural network understand the nature of different features and their interplay. The hierarchical structure of neural networks means the knowledge in each layer builds on that contained in the layers below. In a computer vision algorithm, the lower layers learn simple features like edge and colour detection, the middle layers learn more complicated features and representations, and the higher layers learn the specific semantics of the task at hand. The knowledge in the lower and middle layers is non-task-specific. A non-disease-specific dataset can teach the network this knowledge, especially if it contains large data. On the other hand, the knowledge in higher layers is task-specific. It needs to be learned from task-specific data (e.g., COVID-19 data collected in the first weeks of the pandemic in our case). The utility of transfer learning is most pronounced in problems where task-specific data is scarce, such as a novel disease pandemic [30–32].

Analysis of the performance of different healthcare systems during the pandemic illustrated that the best performing systems had high preparedness and resilience and could scale a response effectively and agilely. Healthcare system resilience includes medical technologies [1]. Data science and AI preparedness should be a part of healthcare system resilience, given the potential impact of AI on optimising pandemic response. We envision RapiD_AI to have the potential to be a pandemic preparedness tool that would give healthcare systems the capability to deploy AI models rapidly after early encounters with a novel pathogen.

We suggest the RapiD_AI framework to support healthcare system preparedness and the effective use of AI during a novel disease pandemic. COVID-19 was arguably the first pandemic that witnessed large-scale experimental use of modern machine learning. Pandemics are expected to increase in frequency and scale. Machine learning can save human life and be a revolutionary enabler of healthcare systems and capabilities. To achieve that, we need to consider the lessons learned from the successes and failures of using AI during the COVID-19 pandemic.

All data in our study comes from multiple facilities in a single city, potentially reducing generalisability. Future research could explore the reproducibility of findings using pretrained models across different populations. Moreover, we have only used a small subset of the potentially usable models. More model architectures should be explored. Finally, we explored the application of the RapiD_AI framework on EHR data only. More data modalities (such as biomedical signals and imaging data) should also be explored.

## 4 Methods

### 4.1 Dataset

In our work, we used three datasets extracted from the same population:

1. 𝒟_*H*_: Jan 2016 till Dec 2019. General in-patient cohort.
2. 𝒟_*W1*_: March 2020 till July 2020. COVID-19 patients only. This represents the first wave of the pandemic.
3. 𝒟_*W2*_: August 2020 till June 2021. COVID-19 patients only. This represents the second wave of the pandemic.

These datasets are de-identified data from the Infections in Oxfordshire Research Database (IORD). This dataset has approvals from Research Ethics Committee, Health Research Authority, and Confidentiality Advisory Group. The data is collected from patients admitted to Oxford University Hospitals (OUH) between January 2016 and June 2021. The data include administrative data, vital sign measurements, laboratory test results, and data on the level of oxygen support. Both 𝒟_*W1*_ and 𝒟_*W2*_ datasets include only data of COVID-19 patients, while 𝒟_*H*_ contains historical data of non-COVID-19 patients. For each evaluated approach, a subset of samples was extracted from each dataset to build the dataframe used for experimentation. Each patient has multiple rows representing the observations recorded during the clinical stay. Observations are recorded at irregular intervals reflecting the time when the hospital staff took the clinical measurements. Each data sample is characterised by a 77-dimensional feature vector and a label exhibiting the possible occurrence of respiratory deterioration events within the next 24 hours (in a retrospective manner). The features include demographic characteristics, vital sign measurements, laboratory test results, and the level of oxygen support measured by FiO2. Moreover, our features include synthetic features that reflect the variability of the clinical features (range, mean of a previous 24-h window) in a trailing window of 24 hours. The full list of features is outlined in D4.

Overall, 𝒟_*W1*_ dataset was used to train models in a way that simulated the first wave of the pandemic, i.e., the samples were split cumulatively into partitions, where partition *i* ∈ {1, …, 20} contains data from week one up to week i [figure C]. Models were trained and validated on each partition and tested on the held out 𝒟_*W2*_ dataset. 𝒟_*W1*_ includes 20 weeks (partitions) that represent the first wave of the pandemic. 𝒟_*H*_ dataset was used to pretrain the models on a source task 𝒯_*Source*_ then each of the resulting models was fine-tuned using 𝒟_*W1*_ to solve a target 𝒯_*Target*_ task [figure 2].

### 4.2 Model architectures

A three-layered fully-connected deep neural network (DNN) was trained to predict respiratory deterioration events. This model consists of three dense layers having 308, 231, and 1 hidden nodes or units. The first two layers are followed by ReLU activation function, and the last one or output layer is followed by sigmoid activation function. A dropout rate of 0.25 is used between each dense layer for regularising the training process. The binary cross-entropy is used as an objective or loss function, and Adam, with a fixed learning rate of 0.0001 is used as an optimiser to train the models. The models are trained for 100 epochs, and early stopping is used to store the best-performing weights or parameters on the validation data.

Gradient boosting machines are a set of machine-learning techniques that combine many weak learning models (models that are only slightly better than the random prediction) to create a strong model [33]. Gradient boosting on decision trees is a well-established ensemble machine learning algorithm that uses trees as weak learners. The idea of boosting is based on sequentially building weak estimators. Initially, all data samples have the same weight. However, at successive iterations, the weights of mislabelled training samples by the boosted model at the previous step are increased, while the weights are decreased for the remaining training samples. Hence, as iterations proceed, the influence of difficult samples is increased. Later, it combines the predictions through a weighted majority vote to reduce the bias of the combined classifier. XGBoost [34] is a highly efficient and accurate implementation of gradient boosting trees known to provide state-of-the-art performance on tabular data.

TabNet [15] is a deep learning-based approach for tabular data problems. It uses sequential attention [35] to determine which features to use at each decision step (i.e., performing instance-wise feature selection), resulting in better interpretability and more efficient learning. The model architecture is composed of multiple steps, each step plays the role of a separate classifier, and they vote together to get the final classification. However, these classifiers are not completely independent since some layers are shared between_steps to provide robust learning and high learning capacity.

### 4.3 Model development and training

We pretrained DNN and TabNet models on 𝒟_*H*_ dataset, then fine-tuned and validated using 𝒟_*W1*_ and finally tested on 𝒟_*W2*_. Since pertaining is not applicable for XGBoost we trained and validated it with 𝒟_*W1*_ and tested it on 𝒟_*W2*_.

For simplicity, we will refer to fine-tuning and XGBoost training as training. During training, we split 𝒟_*W1*_ into cumulative parts, where part i contains data from weeks one up to i. For each part, we applied grid search to find the best HP for each model. Each part is split into train/validation sets (80 / 20). We trained the model on the train set and validated it on the validation set. We optimized the classification threshold using the validation set to get better performance (measured by the AUROC metric). Finally the models were tested on 𝒟_*W2*_.

For XGBoost we tried to tune the number of estimators,(11, 13, 15, 17, 19), their depth (7, 8, 9, 10, 11, 12, 13) and learning rate (0.01, 0.1, 0.2, 0.3). Configuring neural networks is more difficult and sometimes computationally heavy, especially for the TabNet model. So for choosing the architecture and HP, we tend to rely on our understanding of the complexity of our problem and the semantics of the parameters being tuned. For DNN, we used an input layer of size 77, two hidden layers of size 304 and 231, and an output layer of size 1 with a sigmoid activation function. We used a dropout rate of 0.1 between the dense layers and tried to find the optimal learning among [0.0001, 0.0005, 0.001, 0.005, 0.01]. TabNet was the heaviest computationally. We tried the values 32, 64, 128 for n_a and n_d, 5, 7, 9 for n_steps, and 1.5, 1.75 for gamma, however for pretraining, we used n_steps = 5, n_a = n_d = 128 so that our model can generalise to the large number of samples we have.

Among the tested configurations, we used the best parameters set from the first three weeks as the final parameters for our models. After that, we evaluated the models’ stability through confidence intervals. To do that, we used 5-fold cross-validation, and for each split, we tested the model performance on the test set 𝒟_*W2*_. This resulted in 5 measures for the metric, which were then used to find the confidence interval.

After training the models, we calculated the mean performance and the confidence intervals in terms of accuracy, sensitivity, specificity, precision, and AUROC.

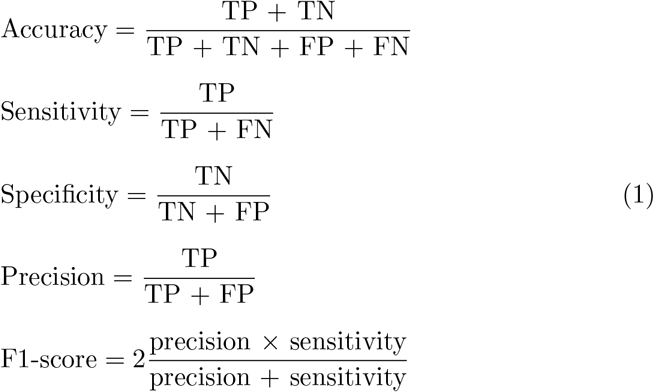

TP, TN, FP, and FN stand for true positive, true negative, false positive, and false negative, respectively. Considering a probability estimate as the output of each classifier for the validation set and setting various thresholds to categorize this output as event/non-event could result in different TP, FP, FN, and TN rates. Alternatively, a receiver operating characteristic (ROC) curve shows the sensitivity as a function of 1 - specificity for different thresholds; each point in the curve indicates a specific value for sensitivity, specificity, and accuracy. AUROC is the area under the ROC curve.

### 4.4 Comparative methods

In this work, we investigated three approaches for our framework. We will refer to these approaches as Scenarios A, B, and C.

In ***Scenario A*** [figure 2 (b1)], we tested a domain adaptation approach to model development. In this approach, a model is trained on source data with a unique distribution 𝒟_*S*_ to solve a task T. We use the resulting weights as initial weights and train it again on target data with different distribution 𝒟_*T*_ to solve the same task T.

We pretrained the DNN and TabNet models to solve the respiratory deterioration task 𝒯_*RD*_ using general population data (i.e., non-COVID-19 patients). We then fine-tuned it with the COVID-19 patient population to solve the same task (𝒯_*RD*_). COVID-19 patients and non-COVID-19 patients represent different domains, and our model aims to benefit from the knowledge gained from the source domain (non-COVID-19 patients) to solve the same task in the target domain (COVID-19 patients).

The stability of this method was examined by extending the pandemic simulation for a longer time. In details, we extended 𝒟_*W1*_ to cover admissions up to January 2021 using COVID-19 patients from 𝒟_*W2*_. The resulting training set (𝒟_*W1*_-extended) contained 2064 patients (60572 samples) while keeping 207 patients (17140 samples) of 𝒟_*W2*_ for testing (𝒟 _*W2*_-truncated).

In this scenario, we hypothesize that we can pretrain effectively using only the most relevant examples from the available historical data. In the current context, the historical examples that demonstrate maximum similarity to the COVID-19 data are regarded as the most relevant. We used K-means clustering and Euclidean distance to select these most relevant examples. K-means clustering is employed to obtain distinct 256 clusters in the historical data. Each cluster or cluster center can be seen as a pseudo-phenotype.

When the COVID-19 data arrives, we compute the Euclidean distance between each COVID-19 example and each cluster centre. We assign a cluster to each COVID-19 example based on the lowest Euclidean distance. In the end, we compute the frequency of each cluster selected by the COVID-19 examples. The clusters with larger selection frequencies are regarded as similar to COVID-19, and their data points are relevant to COVID. By controlling the number of selected clusters, we can control the amount of historical data used for pretraining. In our experiments, we used 10, 20, 30, 40, and 50% of clusters for pretraining. Algorithm 1 outlines the implementation details of this distillation process in detail.

In the last scenario, ***scenario C*** [figure 2 (b3)], we demonstrate how to use pretrained deployed models to build an algorithm that solves a different problem. To do this, we used another useful tool in machine learning called inductive transfer learning [36].

Inductive transfer learning is a type of transfer learning where the source and target domain tasks are different. This approach allows us to use deployed models for some task (source task 𝒯_*Source*_) to rapidly develop a working model for a new (target task 𝒯_*Target*_). Inductive transfer learning helps transfer knowledge from the source task to the target task [36]. Inductive transfer allows us to leverage various models pretrained on different tasks without the need to access the pretraining data itself. To test the value of this approach, we pretrained a model on a ***general deterioration prediction task* 𝒯**_***GD***_ (source task 𝒯_*Source*_) using 𝒟_*H*_ (source data), and then fine-tuned it to solve a ***respiratory deterioration prediction task* 𝒯**_***RD***_ in COVID-19 population (target task 𝒯_*Target*_) using 𝒟_*W1*_ (Target data).

## Data Availability

All data produced in the present study are available upon reasonable request to the authors

## 5 Acknowledgements

## Patient and public involvement

This work uses patients’ data collected by the UK’s National Health Service as part of their care and support. We thank all the people of Oxfordshire who contribute to the Infections in Oxfordshire Research Database. Research Database Team: L Butcher, H Boseley, C Crichton, DW Crook, DW Eyre, O Freeman, J Gearing (community), R Harrington, K Jeffery, M Landray, A Pal, TEA Peto, TP Quan, J Robinson (community), J Sellors, B Shine, AS Walker, D Waller. Patient and Public Panel: G Blower, C Mancey, P McLoughlin, B Nichols.

## Code availability

The source code of this work can be downloaded from https://github.com/AlexYoussef/RapiD_AI

## Appendix A Respiratory Deterioration Event

The level of respiratory support is defined based on the oxygen delivery devices. A list of the devices and the corresponding level of support is documented in Table A1.

**Table A1.** Different respiratory support levels and the corresponding oxygen support devices used at each level.

| Support Level | Devices |
| --- | --- |
| Level 0 | No support required |
| Level 1 | Simple Mask, Venturi Face Mask, Nasal Cannulae, Nebuliser Mask, Oxymask, Tracheostomy Mask |
| Level 2 | Reservoir Mask, Non-rebreather mask |
| Level 3 | High Flow, Non-invasive system, Continuous positive airway pressure (CPAP),AIRVO |

## Appendix B Clusters similar to COVID-19

Table B2 shows the set of disease clusters identified by a physician as similar to COVID-19. Each cluster is presented along with detailed ICD-10 codes.

**Table B2.** ICD-10 codes used in identified clusters Cluster ICD codes

| Cluster | ICD codes |
| --- | --- |
| Viral pneumonia | J09, J100, J101, J108, J110, J111, J118, J120, J121, J122, J123, J128, J129 |
| All causes pneumonia | J13, J14, J150, J151, J152, J153, J154, J155, J156, J157, J158, J159, K160, J168, J170, J171, J172, J173, J178, J180, J181, J182, J188, J189 |
| Asthma | J441, J448, J449, J450, J451, J458, J459 |
| Heart failure | I500, I501, I509, I510, I511, I512, I513, I514, I515, I516, I517, I518, I519, I528 |
| Acute pancreatitis | K850, K851, K852, K853, K858, K859 |

**Table B3.** Top 10 most common ICD-10 codes in the selected historical data clusters

| ICD code | Description |
| --- | --- |
| I10 | Essential (primary) hypertension |
| Z922 | Personal history of drug therapy |
| Z864 | Personal history of psychoactive substance abuse |
| Z501 | Other physical therapy |
| I489 | Unspecified atrial fibrillation and atrial flutter |
| N179 | Acute kidney failure, unspecified |
| Z867 | Personal history of diseases of the circulatory system |
| Z921 | Cardiac resynchronization therapy (CRT) pacemaker |
| E119 | Type 2 diabetes mellitus without complications |
| N390 | Urinary tract infection, site not specified |

## Appendix C COVID-19 pandemic simulation

Figure C shows the process we used to simulate the pandemic; each batch is split into train and validation sets to train the models, and a final test is done on a separate dataset.

**Fig. C1.**
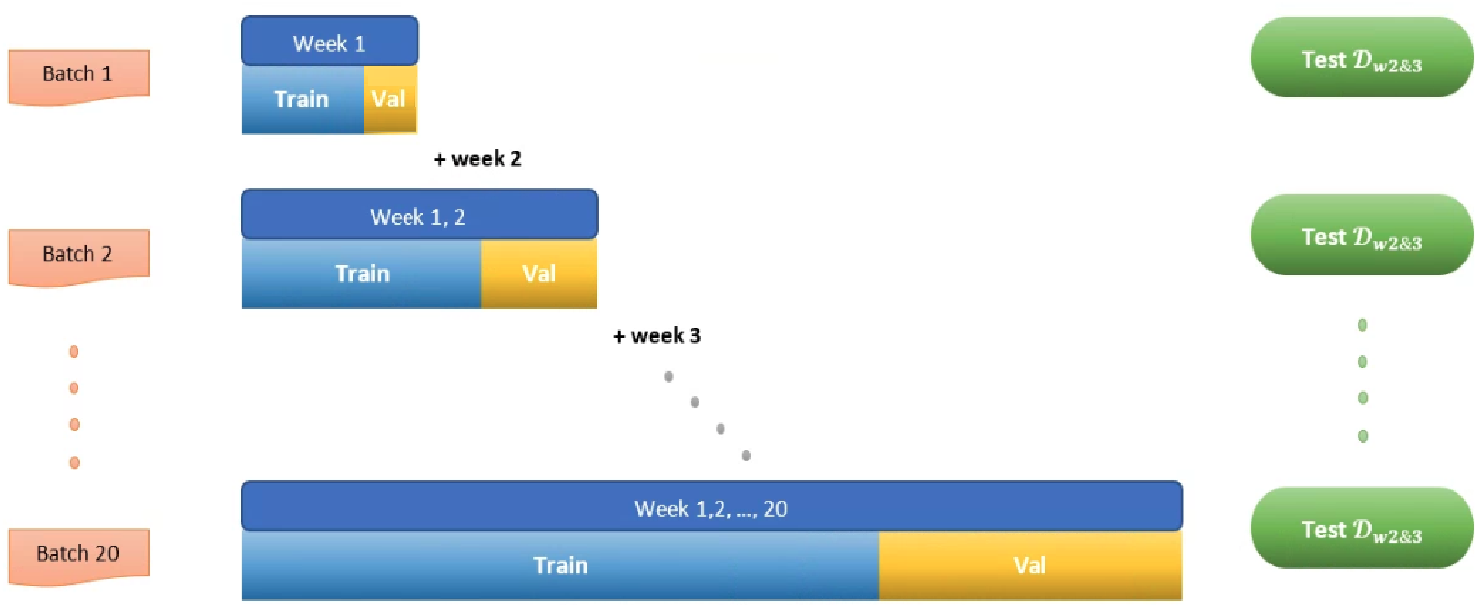
Simulation of COVID-19 pandemic

## Appendix D Features set

Table D4 demonstrates the features used in training our model.

**Table D4.**
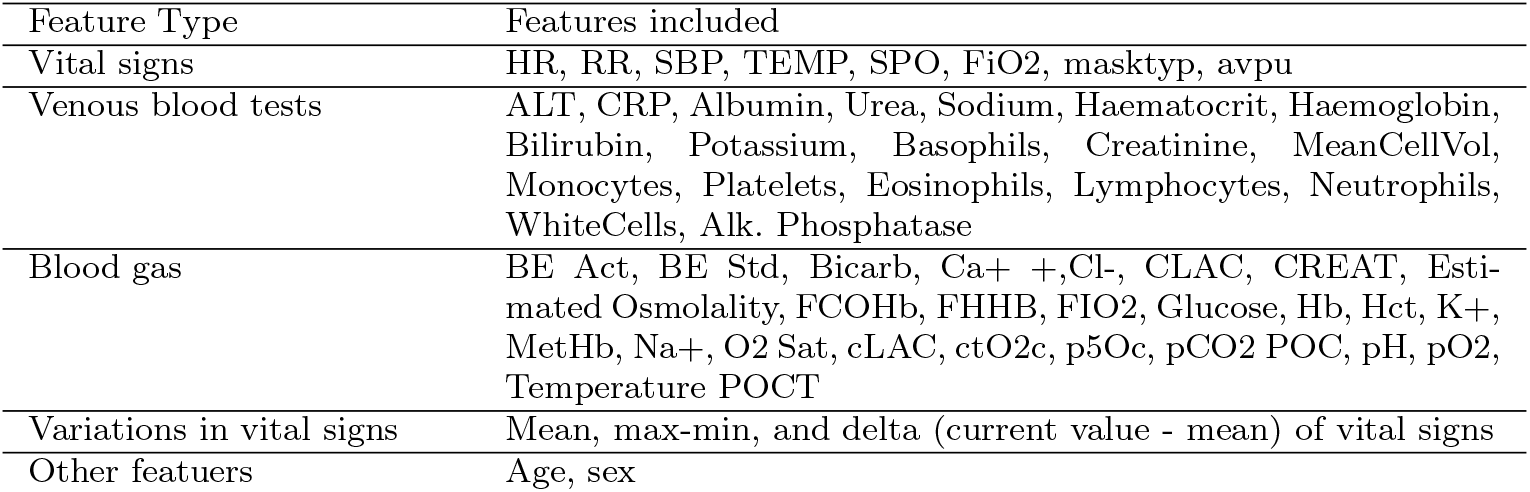
Features included in our extracted datasets

## Appendix E Clustering algorithm

### Algorithm 1 Algorithm to select a subset of historical data for model training

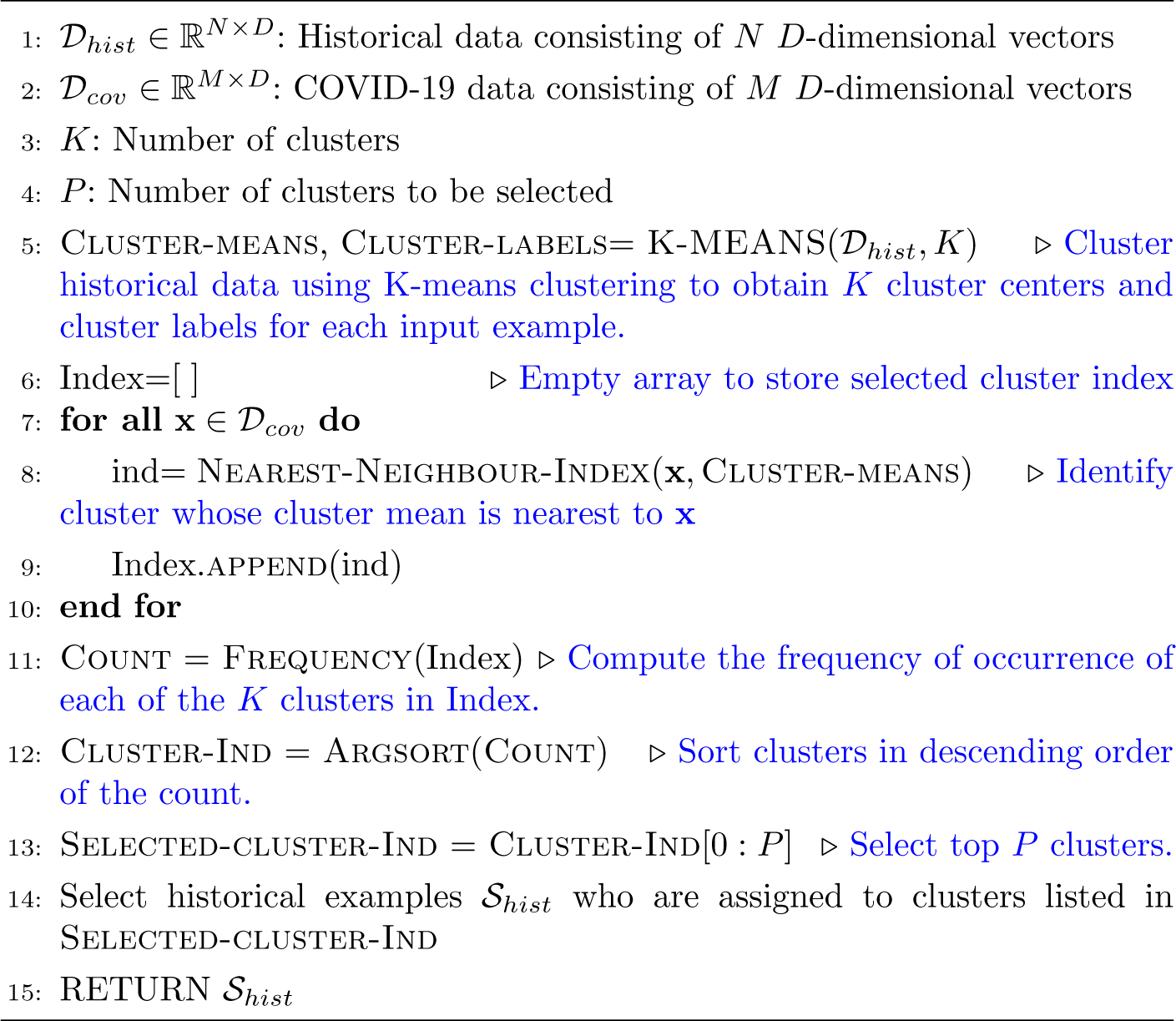

